# The effect of mentorship as a means of strengthening leadership in the health system at the operational level: a case study of the Walungu rural health zone in the eastern Democratic Republic of Congo

**DOI:** 10.1101/2024.05.28.24308067

**Authors:** Rosine Bigirinama, Ghislain Bisimwa, Samuel Makali, Aimé Cikomola, Janvier Barhobagayana, Jean-Corneille Lembebu, Christian Chiribagula, Pacifique Mwene-Batu, Abdon Mukalay, Denis Porignon, Albert Tambwe

## Abstract

**Context:** In the rural Health Zone (HZ) of Walungu, eastern Democratic Republic of Congo, major constraints impede health outcomes. From 2015 to 2019, the “RIPSEC” program transformed Walungu into a Learning and Research Zone (LRZ) under the mentorship of a local university to enhance the leadership capabilities of HZ managers, focusing on managing challenges including the proliferation of Informal Healthcare Facilities (IHFs).

**Objective:** This study evaluates the impact of RIPSEC mentorship on leadership development and the performance of the Walungu LRZ, particularly concerning the utilization of integrated curative health services in competition with IHFs.

**Methods:** We used a mixed method approach, combining retrospective analysis of some key health indicators before (2014) and during RIPSEC program (2015-2019), and in-depth qualitative interviews with members of the HZ management team. Quantitative data were presented as frequencies and proportions. Simple linear regression (p<0.05) measured the influence of IHFs on service use. The functionality and performance of the HZ were assessed using an internal benchmarking approach, with results presented as trend curves. Deductive analysis of interviews allowed for a deeper exploration of quantitative trends.

**Results:** Despite efforts to manage them, IHFs negatively influenced the use of curative services. RIPSEC mentorship notably enhanced the leadership skills of managers, leading to improved management effectiveness. While the use of curative health services slightly increased during the program, rates remained below 50%, and gains were not sustained post-program.

**Conclusion:** RIPSEC mentorship has positively impacted leadership and performance in Walungu. However, challenges related to sustainability and financing persist, necessitating comprehensive interventions beyond enhancing managerial leadership solely.

## Introduction

In contexts of prolonged multifaceted crises, healthcare systems face colossal challenges, progressively undermining their ability to provide quality healthcare to the population (1,2). In 2021, 4.5 billion people lacked coverage by essential health services, and 1 billion faced catastrophic healthcare costs, with 344 million people living in extreme poverty due to these expenses. (3). This reality is starkly evident in the east of the Democratic Republic of Congo (DRC), where decades of armed conflict have fostered a persistent crisis (4). The ongoing conflict has gradually eroded infrastructure and led to an inadequate distribution of healthcare resources (1), while the state struggles to finance quality, accessible health services for all. With less than 15% of the national budget allocated to health, and only 30% of this amount disbursed, the majority of healthcare financing falls on households, which contribute 40% of the overall health budget, 90% of which is through direct payments (5–8). As a result, health facilities commonly rely on direct payment for care by patients to ensure their operation (7,8). These financial constraints significantly deter the sick from using healthcare services (9), contributing to the population’s growing impoverishment and poor health outcomes.

Moreover, the limited government presence in crisis zones (10) has led to the proliferation of Informal Healthcare Facilities (IHFs). These facilities operate marginally within legal frameworks, often failing to meet sectoral standards and posing risks to patient safety (11). Despite these issues, economic constraints and often misleading promises draw people to these facilities (12). The inability of the state to effectively regulate these entities is further complicated by political interferences that undermine the regulatory attempts of HZ managers (11,13). However, strengthening Health Zone (HZ) management is recognized as a pivotal strategy for addressing these challenges, as demonstrated in various contexts, including those similar to the east of the DRC (15–18).

In the DRC, the organizational arrangements of the health system are such that leadership must be exercised fully at the lowest level of the health system, through the HZ managers gathered in a Health Zone Managment Team (HZMT) (18). The HZMT is responsible for managing and coordinating health services, planning interventions, conducting epidemiological surveillance, and supporting primary care structures. They also have the task of ensuring the effective implementation of national health policies (18,19). Previous studies in eastern DRC have identified a notable leadership deficit among managers in crisis zones, with no formal government program currently aimed at enhancing HZ management leadership (11,20). In 2015, an evidence-based health system strengthening program named “Renforcement Institutionnel pour des Politiques de Santé basées sur l’Évidence en RD Congo” (RIPSEC) was established in the Walungu HZ, South-Kivu province, targeting it as a “demonstration health zone,” a strategy proposed in the DRC’s National Health System Strengthening Strategy but not yet implemented (18). Walungu was selected since it met key selection criteria: it presented a minimally acceptable level of functionality, was geographically accessible, and had favorable security conditions. These criteria ensured that the selected zone could provide a safe, stable, and accessible environment for effective HZMT mentoring. Walungu was positioned to serve as a model for other HZs, demonstrating viable strategies for developing and improving health services.

This study is based on the hypothesis that leadership development through the “demonstration health zone” approach can effectively address the challenges in crisis-affected areas. It specifically examines the impact of RIPSEC mentorship on the leadership capabilities of the Walungu HZMT in the face of competition from IHFs.

## Methodology

### Study region: presentation of the case under study

The study was conducted in the Walungu HZ in the South-Kivu province of eastern Democratic Republic of Congo (DRC). Walungu is one of 519 operational HZs (also known as Health Districts) that form the foundational layer of the DRC’s health system pyramid. Each HZ is structured to provide two primary healthcare service lines. The first line consists of a network of 7 to 25 Health Centers (HCs), each serving its corresponding geographical entity, known as a Health Area (HA), and delivering a minimum package of care. The second line is typically represented by a General Referral Hospital (GRH) that offers a complementary package of care. Some HZs also include secondary hospitals or Referral Health Centers that provide a level of care between the first and second lines. The intermediate coordinating level of the health system pyramid comprises 26 Provincial Health Divisions (DPS), one for each of the country’s 26 provinces, while the central normative level is headquartered in Kinshasa, the capital city.

Walungu is characterized as a rural HZ with an estimated population of 285 669 in 2019, distributed across 23 HAs (Table 1). According to the 2020 report from the HZ Central Office, the estimated coverage rate is 96%. This is defined as the population living within less than 5 km from an integrated health facility without significant geographical barriers (19). Similar to most HZs in eastern DRC, Walungu is a post-conflict zone that continues to experience sporadic attacks on its population by armed rebel groups (1,21).

**Table 1.** General characteristics of the Walungu HZ.

|  | Baseline | RIPSEC Program Years |  |  |  |  |
| --- | --- | --- | --- | --- | --- | --- |
|  | 2014 | 2015 | 2016 | 2017 | 2018 | 2019 |
| **Variables** |  |  |  |  |  |  |
| **Population (inhabitants)** | 244684 | 252269 | 260362 | 268434 | 276755 | 285669 |
| **Number HAs** | 23 | 23 | 23 | 23 | 23 | 23 |
| **Number of GRH** | 1 | 1 | 1 | 1 | 1 | 1 |
| **Number Referral Health Centers** | 5 | 5 | 5 | 5 | 5 | 5 |
| **Number of Health Centers** | 23 | 23 | 23 | 23 | 23 | 23 |
| **Number of Health Posts** | 3 | 4 | 6 | 6 | 6 | 7 |
| **Number of beds in the GRH** | 200 | 200 | 235 | 233 | 286 | 272 |
| **Health Coverage** | 96.4% | 96.4% | 96.4% | 96.4% | 96.4% | 96.4% |
| **Number of medical doctors** | 9 | 9 | 9 | 9 | 9 | 10 |
| **Number of nurses** | 121 | 127 | 137 | 164 | 153 | 201 |
| **Medical doctors /10000 inhab.** | 0.37 | 0.36 | 0.35 | 0.34 | 0.33 | 0.35 |
| **Nurses / 10000 inhab.** | 4.9 | 5.0 | 5.3 | 6.1 | 5.5 | 7.0 |
| **Beds / 10000 inhab.** | 8.2 | 7.9 | 9.0 | 8.7 | 10.3 | 9.5 |

### Intervention framework of the RIPSEC program in Walungu

The RIPSEC program, led by a consortium of three Schools of Public Health in the DRC and funded by the European Union, was operational from 2015 to 2019. Its overarching goal was to enhance the use of scientific evidence in health policy development to improve health outcomes. It implemented three LRZs across three provinces, the program led by three main DRC’s academic public health institutions: UNILU in Haut-Katanga province (for Kisanga HZ), UNIKIN in Kongo Central province (for Gombe Matadi HZ), and “Ecole Régionale de Santé Publique” of the “Université Catholique de Bukavu” (ERSP-UCB) in South-Kivu province (for Walungu HZ) (22).

In Walungu, the RIPSEC program included four major “transformation projects” designed to boost the effectiveness of the HZMT. These components were: (i) leadership strengthening, (ii) formative supervision, (iii) enhancement of referral and counter-referral systems, and (iv) transformation of health facilities into model structures for other HZs.

Leadership development was customized to fit the specific needs of Walungu, focusing on enhancing functionality and ensuring efficient coordination of activities.

The mentorship was led by a well-defined profile: a public health expert and experienced former HZ manager who was working, at the time of his selection, as manager of a health program at provincial level. This mentor was responsible for organizing weekly HZMT meetings, providing individual coaching to foster greater commitment and participation in management activities. Formative supervision replaced previous supervisory approaches with interactive, on-the-spot error correction and detailed reporting post-visit. The standardization of referral and counter-referral tools was aimed at harmonizing procedures and improving communication across facilities. Additionally, facilitating exchanges between lower and higher-performing facilities aimed to enhance performance indicators and disseminate best practices. All activities were meticulously documented under the oversight of ERSP-UCB (23).

### Conceptual framework

The framework (Fig 1) conceptualizes the transformative potential of targeted leadership support within a complex socio-economic, cultural, and administrative context marked by post-conflict recovery. It delineates the process by which leadership enhancement of the HZMT through RIPSEC’s mentorship is expected to directly improve the overall functionality and performance of the HZ. Activities are strategically designed to address the unique challenges encountered in Walungu effectively. Expected outcomes include an increased utilization of curative services and a notable reduction in the negative impacts of IHFs, among other potential improvements.

**Fig 1.**
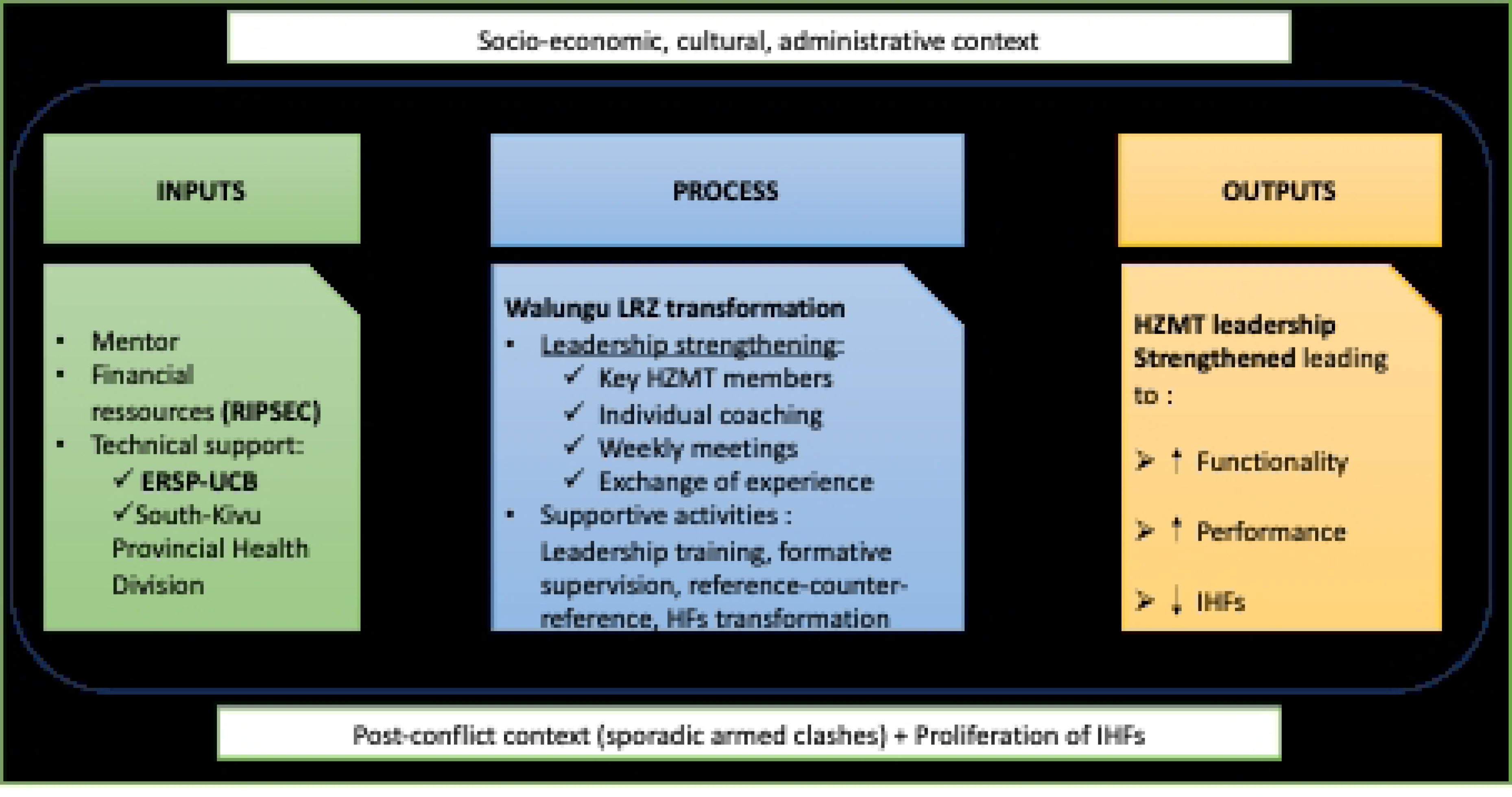
Conceptual framework of the RIPSEC program intervention in Walungu LRZ.

### Definition of concepts

1. Mentorship: In the context of public health, mentorship refers to a supportive and advisory relationship where an experienced individual (the mentor) guides and assists a less experienced individual (the mentee) in their professional development. The goal is to enhance the mentee’s skills, knowledge, and confidence (22). Within the RIPSEC program, a public health expert from the ERSP-UCB was assigned to provide ongoing technical support, training, advice, and local coaching to improve leadership and management skills of the HZ managers (22,23).
2. Learning and Research Zone (LRZ): An LRZ is a designated environment that facilitates learning, research, and the development of improved practices through collaboration between researchers and health managers (13,24). In the RIPSEC context, the Walungu LRZ served as a testbed for innovative strategies and interventions aimed at enhancing the healthcare system and generating evidence. The mentorship activities described in this study are integral components of this LRZ’s function (22).
3. Health structures: Walungu HZ encompasses a range of health facilities (HF) offering diverse healthcare needs. These include:
  - Health Centers: First-line HF that deliver a minimum package of services, including curative, preventive, promotional, and rehabilitation activities.
  - Walungu General Referral Hospital: A second-line medical facility that offers a complementary package of activities, including the four essential services of Internal Medicine, Surgery, Gynaeco-obstetrics, and Pediatrics.
  Additionally, the zone features various other health structures that provide a spectrum of care levels. At one end are Health Posts, offering the most basic care package, and at the other, Reference Health Centers equipped with maternity units and comprehensive medical consultation services, going beyond the minimum care package (19). Among these structures, some are integrated. Integrated structures are state-approved facilities adhering to national health standards and reporting to the Ministry of Health. Conversely, private facilities, though state-approved, are not mandated to report their data to the Ministry of Health(18,25).
4. Informal Healthcare Facilities (IHFs) : These are entities that purport to offer care aimed at curing or alleviating illnesses without the requisite quality or authorization. They fall into two categories:

- Traditional Medicine facilities: Structures claiming to treat illnesses using traditional methods and materials such as plants, leaves, roots, and animal parts not recognized in modern pharmacology.
- Prayer-based facilities: Facilities that organize prayer sessions or vigils seeking divine healing.

### Type and period of study

This study is an analysis of the implementation of the RIPSEC program in the Walungu LRZ, employing a single case study approach. We employed a mixed-methods approach for exploratory purposes (Table 2), combining retrospective analysis of key health indicators from 2014 (pre-implementation) through the five-year duration of the RIPSEC project (2015–2019), with in-depth qualitative interviews conducted post-program. Data triangulation from these approaches was used to enhance the robustness of our findings.(27–29). Quantitative data were collected continuously from January 1^st^, 2015, to December 23^rd^, 2019, and were accessed for analysis on March 23^rd^, 2022. The qualitative component, focusing on the program’s contribution to leadership and governance development, involved interviews with key HZMT members. These interviews were conducted from November 23^rd^ to December 15^th^, 2022, to assess the enduring impacts and gather reflective insights from the stakeholders involved.

**Table 2.**
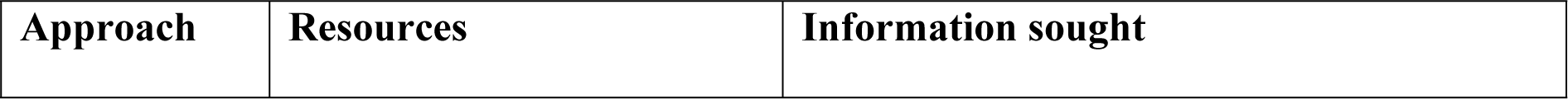
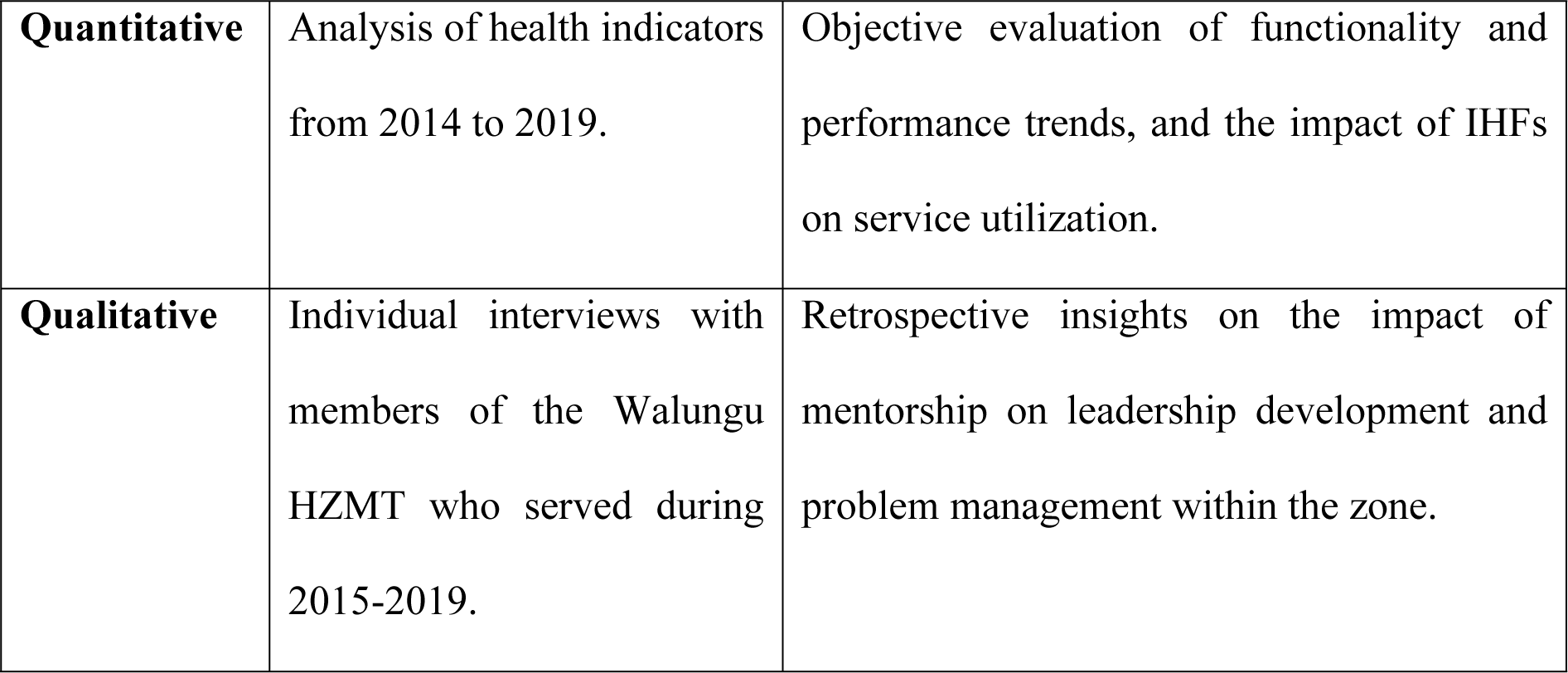
Summary of study approaches.

### Sampling and data collection

#### 1) Quantitative component of the study

To assess the evolution of the zone’s functionality and performance, exhaustive sampling was employed, focusing on key health indicators at the HZ, particularly those related to service utilization in integrated facilities. During the RIPSEC program period, deidentified data were gathered from secondary sources, specifically health reports at both the HF and HZ Central Office levels, by the program mentor. This data collection process ensured that there was no access to individual identities during and after data collection. The mentor, serving as a part-time technical assistant, played a pivotal role in organizing the HZ, collecting data monthly, and coaching the HZMT. The mentor’s presence during the RIPSEC program ensured the reliability of the data recorded in monthly reports. For observations prior to the LRZ establishment, the scope of the data collection was restricted to the year 2014. This limitation was due to the incomplete implementation of the national DHIS2 data system at that time (29) and the absence of the RIPSEC mentor in the zone. Additionally, archival weaknesses during this period further restricted the reliability of any data prior to 2014. Collected data were stored on a secured disk at the ERSP-UCB.

Performance indicators were specifically selected to reflect preventive activities at first-line HFs and curative activities at second-line HFs. This study emphasized the rate of use of curative services in Walungu’s HFs as a proxy for assessing the population’s reliance on IHFs: a lower usage rate suggests a higher likelihood of the population turning to alternative curative options. These indicators, presented as rates, were chosen to mirror the impact of managerial leadership in health service delivery (4,14) (Table 3).

**Table 3.**
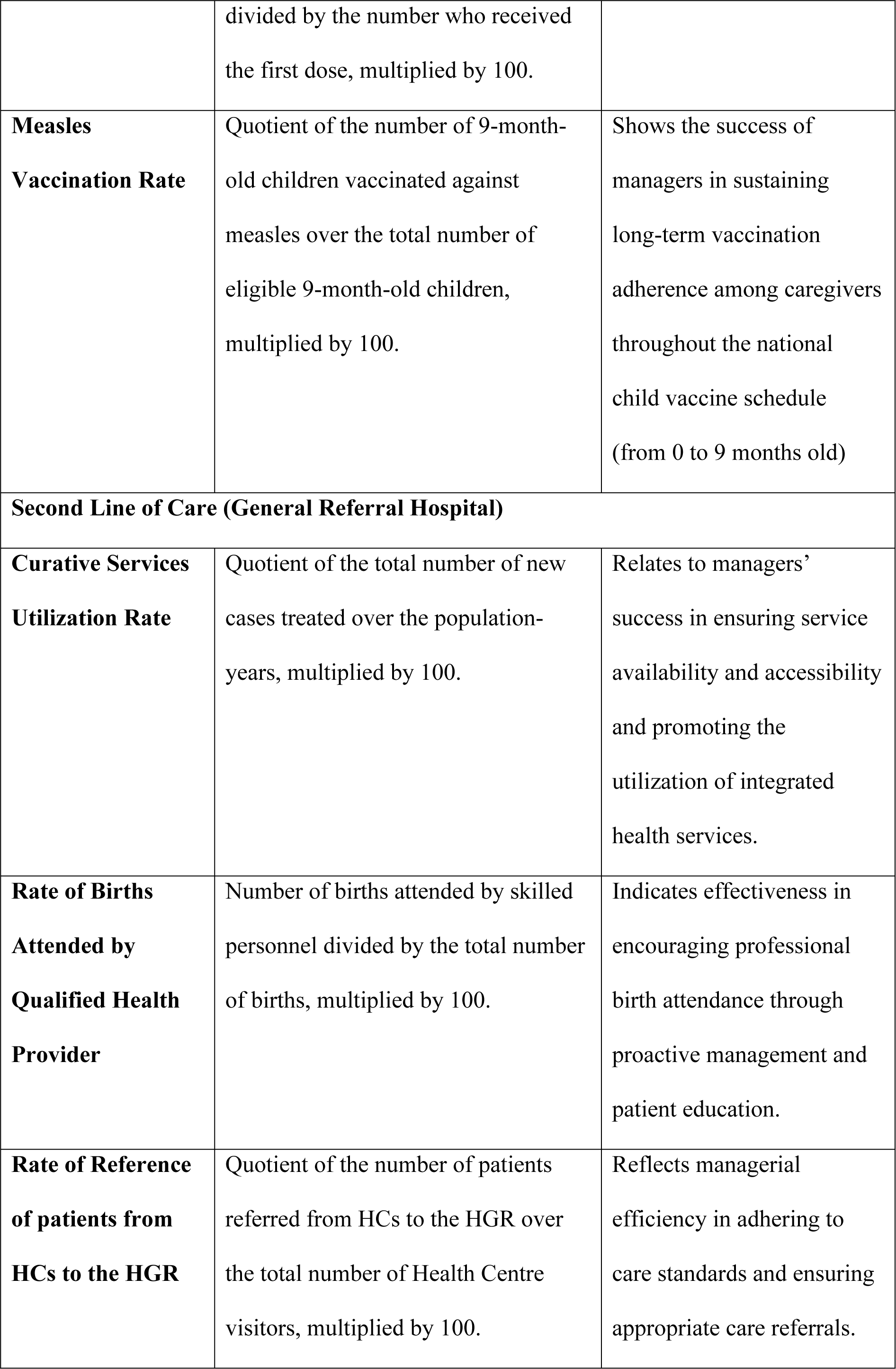
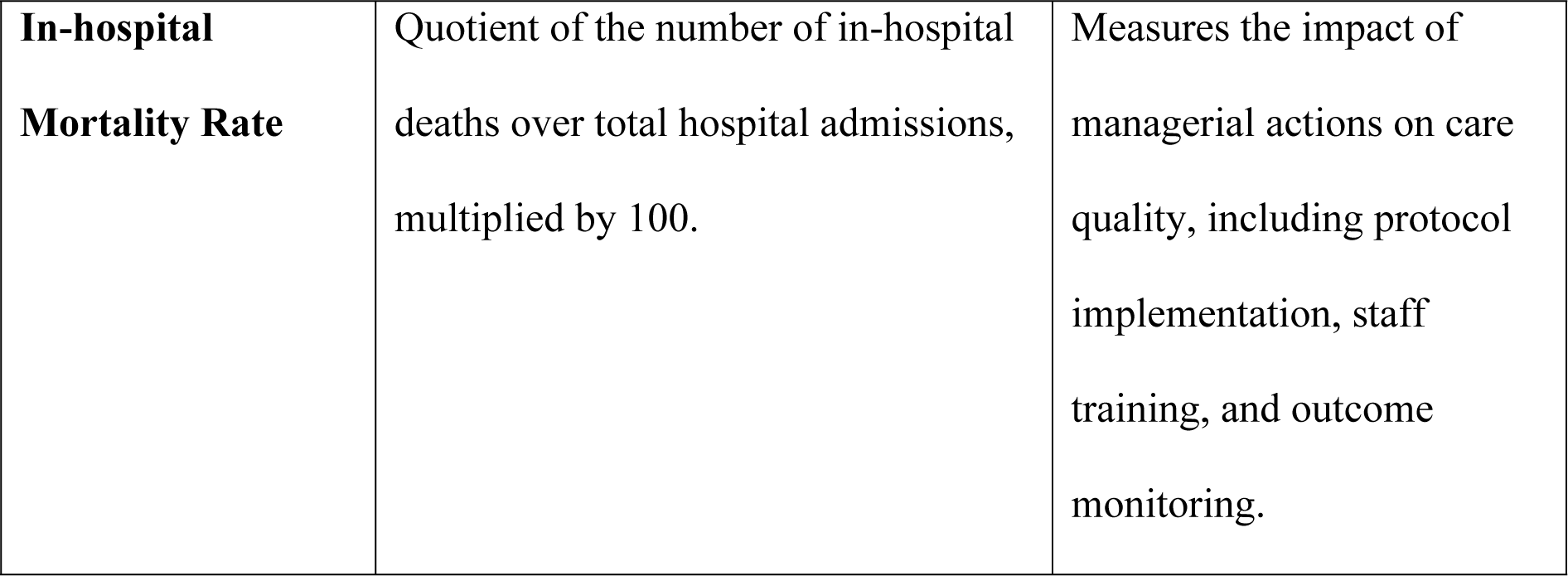
Performance indicator.

| Indicator | Definition | Link with Leadership |
| --- | --- | --- |
| **First Line of Care (Health Centre)** |  |  |
| **First Antenatal Visit Utilization rate** | Quotient of the number of pregnant women who attended their first antenatal visit over the total number of eligible pregnant women in the HZ, multiplied by 100. | Reflects the effectiveness of HZ managers in promoting initial prenatal consultations among pregnant women. |
| **Fourth Antenatal Visit Utilization Rate** | Quotient of the number of pregnant women who attended all previous antenatal appointments over the total number of eligible pregnant women in the HZ, multiplied by 100. | Demonstrates the ability of managers to ensure continued engagement of pregnant women in ongoing prenatal care. |
| **Third Dose Pentavalent Vaccine Wastage Rate** | Difference between the number of children who received the first and third doses of the pentavalent vaccine, | Indicates managerial effectiveness in maintaining adherence to the national child vaccination schedule. |
|  | divided by the number who received the first dose, multiplied by 100. |  |
| **Measles Vaccination Rate** | Quotient of the number of 9-month-old children vaccinated against measles over the total number of eligible 9-month-old children, multiplied by 100. | Shows the success of managers in sustaining long-term vaccination adherence among caregivers throughout the national child vaccine schedule (from 0 to 9 months old) |
| **Second Line of Care (General Referral Hospital)** |  |  |
| **Curative Services Utilization Rate** | Quotient of the total number of new cases treated over the population-years, multiplied by 100. | Relates to managers' success in ensuring service availability and accessibility and promoting the utilization of integrated health services. |
| **Rate of Births Attended by Qualified Health Provider** | Number of births attended by skilled personnel divided by the total number of births, multiplied by 100. | Indicates effectiveness in encouraging professional birth attendance through proactive management and patient education. |
| **Rate of Reference of patients from HCs to the HGR** | Quotient of the number of patients referred from HCs to the HGR over the total number of Health Centre visitors, multiplied by 100. | Reflects managerial efficiency in adhering to care standards and ensuring appropriate care referrals. |
| **In-hospital Mortality Rate** | Quotient of the number of in-hospital deaths over total hospital admissions, multiplied by 100. | Measures the impact of managerial actions on care quality, including protocol implementation, staff training, and outcome monitoring. |

For functionality, indicators aligned with leadership and good governance were chosen. These include organizational and managerial capacities typically found in a HZ manager demonstrating effective leadership. Such indicators include the frequency of various meetings, such as the Board of Directors meeting (encompassing HZMT, provincial-level managers, and technical partners from the zone), Management Board meeting (HZ Central Office and GRH managers), HZ Central Office Board of Directors meeting, GRH Board of Directors meeting, HZ Reviews, and Health Development Committee of the HZ meetings (14).

To understand the dynamics of the population using IHFs, including traditional medicine and prayer groups, we employed an indirect indicator— the number of IHF providers identified in the HAs. We posited that the proliferation of IHFs incentivizes their use (12); thus, the higher the number of IHFs in a HA, the greater the impact on curative services rates.

We categorized Walungu’s HAs into two groups based on the average number of IHFs: “safe HAs” were those HAs counting less than the average number of IHFs per HA in Walungu, and “infiltrated HAs” were those counting at least the average number of IHFs within them. Thus, of the 23 HAs in the zone, 10 have been categorized as “safe HAs” and 13 as “infiltrated HAs”. This data has been available since 2014 and was assumed to be stable through 2019.

#### 2) Qualitative component

In-depth interviews were conducted with Walungu HZMT members who had served for at least two consecutive years during or overlapping with the RIPSEC program period. Members eligible for the study were identified from the RIPSEC archives and subsequently contacted. Seven members, all male, agreed to participate. The interviews were carried out either face-to-face or remotely, depending on the participants’ availability, and verbal informed consent was obtained prior to each session. Each interview lasted about 43 minutes. The interview guide was developed based on the preliminary analysis of quantitative data, and the interviews were conducted by two senior researchers with extensive experience in qualitative methods. All interview audios and transcripts were securely stored on a disk at the ERSP-UCB.

##### Note

While the contribution of various technical and financial partners is acknowledged as vital in strengthening the DRC’s health system, often described as essential to the basic functioning of the majority of the region’s HZs (30), this study deliberately narrows its focus to the specific impact of the RIPSEC program. This focused approach is justified by the unique nature of the RIPSEC intervention, which integrates mentorship and leadership development, components not typically emphasized to the same extent in other partnerships. By isolating the effects of RIPSEC, we aim to assess the distinct influence of its mentorship and training components on the functionality and performance of Walungu. Moreover, treating the interventions of other partners as elements of the overall context allows for a more controlled study of RIPSEC’s direct contributions, providing valuable insights into the effectiveness of targeted mentorship and leadership training in health system strengthening. This methodological choice enhances the study’s specificity and relevance, for a more precise understanding of how focused leadership initiatives can impact health outcomes in crisis-affected areas.

### Analysis

The data collected in HFs were organized on MS-Excel spreadsheets, and analyzed using MS-Excel for some, and STATA Version 17 for others. Simple linear regression was used to assess the influence of IHF presence on curative service utilization in 2014 and during the LRZ years (2015–2019), with significance set at p<0.05.

Functionality and performance were evaluated using an internal benchmarking method (31,32), starting with indicator selection, and scoring (ranging from 0 to 4, where 4 indicates the target achieved). Scores were adjusted based on expected results and predefined standards, allowing for comparative analysis. Walungu’s performance in the 13 selected indicators was benchmarked comparatively to standards used by the DRC’s health system to assess HZs achievements (4,31,33). Results were illustrated through curves showing changes from 2014 to 2019.

For the qualitative data, a deductive thematic analysis was applied to the interview transcripts, which were anonymized and coded from RP1 to RP7. The analysis framework was based on the interview guide, enriched by the initial quantitative findings. Themes included (i) the mentorship’s contribution to leadership, (ii) its effect on HZ functionality and performance, and particularly, (iii) its effect on curative service usage confronted to IHFs. Manual coding identified relevant categories, and thematic grouping facilitated theory development and sub-theme identification.

### Ethical considerations

The research protocol for this study was approved by the ethics committee of the Université Catholique de Bukavu under the reference number UCB/CIES/NC/019/2021. The approval was granted under the title “Support for Strengthening the Management and Leadership Skills of the National Malaria Control Program Management Team”. All stages of the study complied with the relevant guidelines and regulations. All participants provided informed consent before taking part in the study.

## Results

### The use of curative services before and during RIPSEC program

We observed an improvement in the use of curative care services in Walungu’s safe HAs from the LRZ years onwards (Fig 2), while there was a deterioration in infiltrated HAs over the same period. Safe HAs consistently showed better rates of curative service utilization. In 2018, there was a sharp decline in the rates for both types of HAs, which subsequently recovered in 2019. It is important to note that the utilization rates for curative services remained below 50% across all HAs at all times.

**Fig 2.**
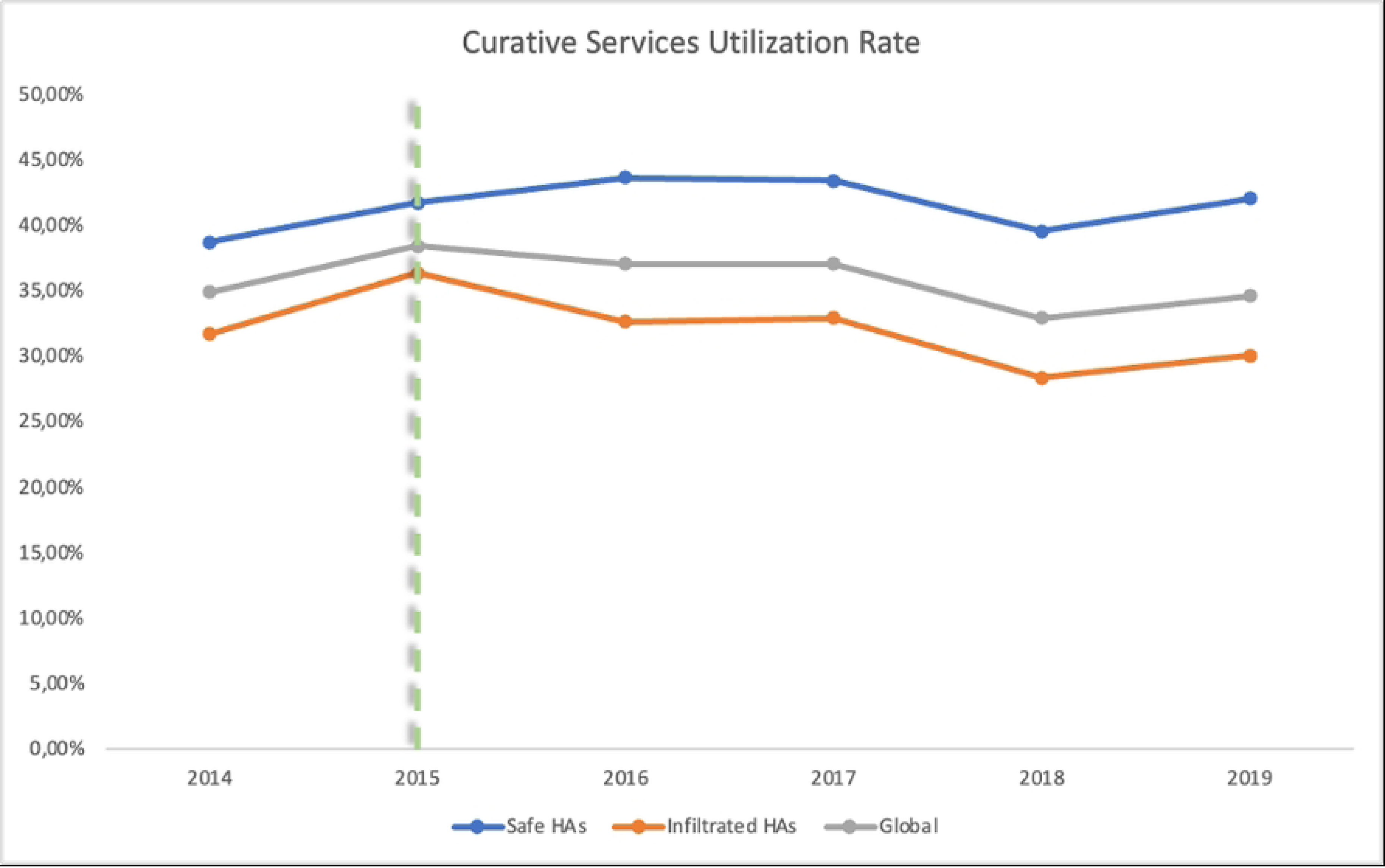
Curative healthcare services utilization rate trends (the dotted line delimits the periods before and during RIPSEC)

### Influence of IHFs on the use of curative services

Table 4 presents the results from a simple linear regression that assesses the influence of IHFs on the rate of use of curative services in Walungu’s HAs. The β value for safe HAs before RIPSEC indicated a trend towards higher use of curative services compared to infiltrated HAs, although this difference was not statistically significant (p=0.062). During the mentorship years, safe HAs had a statistically significant higher average rate of use of curative services (β=0.10, p=0.007) compared to infiltrated HAs. These findings suggest that the presence of IHFs negatively influences the utilization of curative services.

**Table 4.** Use of curative services and IHFs before and during RIPSEC program.

| Variables | Before RIPSEC (2014) |  | During RIPSEC (2015-2019) |  |
| --- | --- | --- | --- | --- |
| | $\beta$ (95% CI) | p-value | $\beta$ (95% CI) | p-value |
| **Types of HAs** |  |  |  |  |
| **Safe HAs** | 0.07 (-.001 ; 0.13) | 0,062 | 0.10 (0.03 ; 0.17) | 0.007 |
| **Infiltrated HAs** | 0 |  | 0 |  |

### Walungu’s achievements before and during RIPSEC program

An analysis of trends in the selected indicators is summarized in the figures above. Generally, both curative and preventive activities exhibit positive trends, with preventive activities consistently recording better performance scores than curative ones (Fig 3). Regarding the management indicators, there is an overall upward trend; however, a distinct decline occurred in 2018, followed by a recovery. The analysis of the evolution of functionality and performance (Fig 4) indicates an overall improvement during the RIPSEC years compared to 2014. Notably, performance consistently exceeds functionality across the years, with the same sharp decline in 2018 as previously observed.

**Fig 3.**
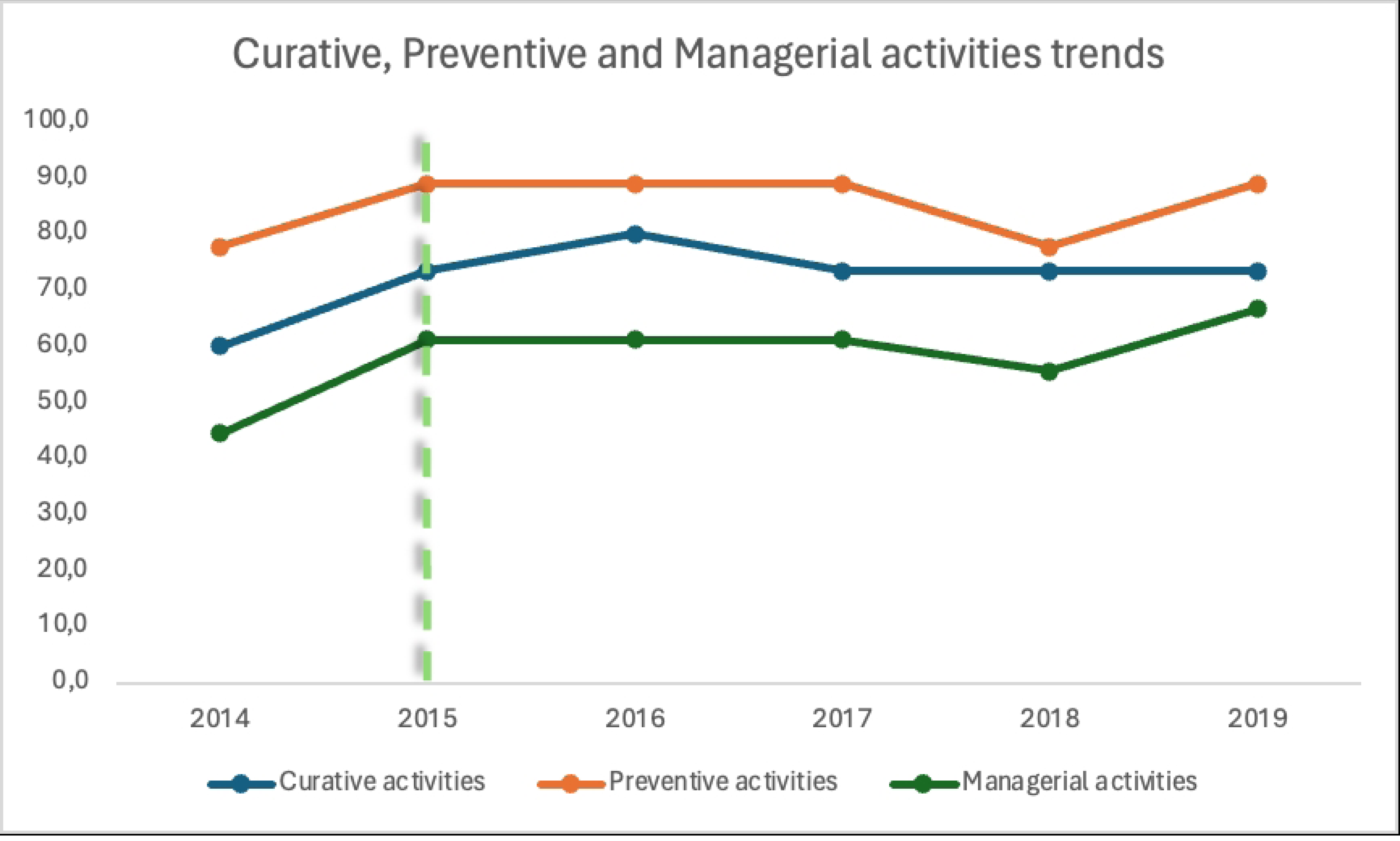
Preventive, curative and managerial activities trends in Walungu (the dotted line delimits the periods before and during RIPSEC)

**Fig 4.**
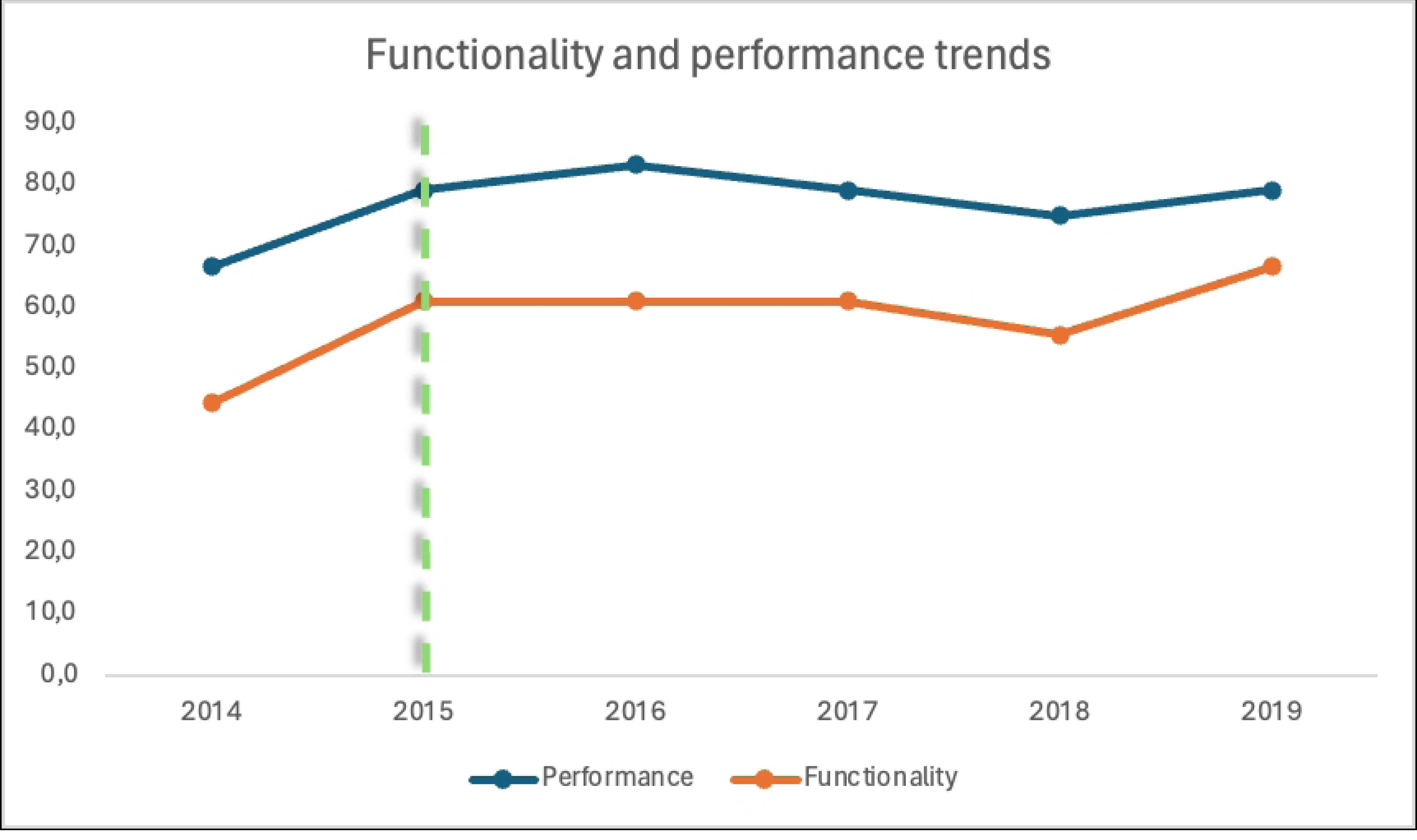
Functionality and performance trends in Walungu (the dotted line delimits the periods before and during RIPSEC)

### HZMT members’ experience of RIPSEC mentorship

Our thematic analysis of the interview transcripts deduced three primary themes, with a fourth unexpected theme emerging: the challenges encountered in sustaining the achievements of the RIPSEC program.

### Leadership under RIPSEC mentorship

The interviews highlighted a strengthening of leadership within the HZ management during the program, emphasizing the significance of a clear vision and effective decision-making. Participants frequently credited the RIPSEC program as a catalyst for this transformation by introducing crucial leadership training and strategies to address identified gaps. Despite financial constraints and limited resources, the training and capacity-building initiatives positively impacted care quality and patient management. Respondents noted improvements in accountability, transparency, and coordination across different health management levels.

> “Leadership has really changed the way we run services… now there is more accountability and follow-through” (RP1).

> “Leadership is crucial. Before RIPSEC leadership training, our efforts were scattered.” (RP5)

> “We needed leaders who understood our challenges and could guide effectively.” (RP3)

### Improving health outcomes in the area

Respondents recognized staff training and skills development as positive aspects of the RIPSEC program. These initiatives have helped stabilize health indicators in accordance with standards. Specifically, nurse managers at HCs were trained to enhance quality of care within their facilities, which improved clinical skills and positively affected health indicators, case management, and team communication through frequent capacity sharing sessions.

> “The training courses have greatly helped our staff to manage cases more effectively” (RP5).

> “Communication mechanisms have been put in place to enable nurses and referred patients to make contact with the doctors who will see patients in hospital.” (RP3)

> “I can personally say that through the activities that have been carried out with the community, my skills and knowledge have increased as a result of sharing experiences with others. Although I wasn’t directly involved, but with the exchanges of experience that we did, it was beneficial for everyone, for the community.” (RP7)

### Use of curative services in competition from IHFs

Participants discussed how informal care structures, such as prayer houses and traditional healers, significantly attract the population, impacting the use of formal health services, especially in geographically hard to reach HAs. Cultural beliefs and socio-economic challenges as well as poor governance practices have hindered integration of these facilities into a regulated framework. The RIPSEC mentorship attempted to address these issues by developing several initiatives to curb this unregulated competition, leading to improved coordination of care and increased health awareness.

> “When you arrive in almost all the zones at the moment, it’s the people from X territory who have unlicensed structures because they currently have a brother from the same territory who is a minister. Now he’s encouraged them to open pirate facilities and you, as the zone’s chief medical officer, have no power over them. “(RP1)

> “Within the RIPSEC project, we sought to involve local authorities with the aim of raising awareness of the need to monitor informal care homes.” (RP4)

> “With the presence of these prayer rooms, many health facilities are not able to attract as many patients as they should… We have identified and drawn up a list of these problematic facilities in collaboration with community leaders.” (RP6)

> “RIPSEC had trained and informed the community health workers on how to manage and listen carefully to the community’s problems… I saw the community health workers trained by RIPSEC go out into the community to raise awareness or inform people about their rights but also their advantage of attending the integrated health center.” (RP7)

### Sustaining RIPSEC’s achievements

Challenges related to disruptive events were noted. Respondents mentioned, among other things, obstacles such as internal conflicts and the instability of human resources (particularly the 2018 Ebola Virus epidemic, the death of the GRH medical director and the departure of the zone’s chief medical officer, in 2018), which significantly slowed down the project’s progress. Concerns were raised about resources gaps and the long-term sustainability of improvements.

> “Since RIPSEC, we have seen better use of services, but there are still challenges to be overcome” (RP3)

> “The lack of funding and equipment is a major challenge for us.” (RP2)

> “In 2018, first there were staff disruptions at hospital level. And then there was the Ebola epidemic, there were the unfortunate events that followed: there were doctors who died in the hospital in the space of two years, and who had benefited from RIPSEC training.” (RP5)

The limited duration of the program (5 years) was frequently cited as a barrier to achieving lasting success and hindered the transmission of LRZ achievements and good practices to other provincial HZs.

> Providers with experience in managing deliveries at the General Referral Hospital were going to work in the two health centers in order to raise the level (of competence of the providers) in the health centers… RIPSEC stopped when all 4 projects were not yet well covered. If I had the power, I would relaunch RIPSEC. And as I said, we were a pilot health zone, we were supposed to set an example for the other zones, but unfortunately the project came to an end and the other zones stopped coming… (RP1)

## Discussion

### Overview

The aim of this study was to explore the impact of a mentorship-based leadership development program on the functionality and performance of a HZ, and on its capacity to resolve specific problems, in a crisis context. We focused on the Walungu HZ, faced with the specific problem of the presence of IHFs. We investigated how the transformation of the Walungu into a LRZ under the RIPSEC program’s mentorship influenced health outcomes within the HZ. Findings reveal that while the program led to a modest but significant improvement in health service utilization and management capabilities, the persistent influence of IHFs, exacerbated by socio-economic and governance factors, continue to pose significant challenges. Despite the progress made through mentorship, sustainability strategies and continuous support are required to maintain and effectively integrate these initiatives into existing healthcare systems.

### The challenge of IHFs

The rate of service utilization in Walungu remains low, falling below the standard threshold of 50% (19), in stark contrast to the provincial average which increased from 34% to 53% during the same period (5).

Trend curves for the use of curative health services indicate a decline in formal service usage in areas with a high concentration of IHFs, suggesting that regulatory efforts in these areas are less effective. Respondents indicated that influence peddling often places these informal entities beyond legal oversight, undermining the regulatory authority of HZ managers. The limited decision space available to these managers, constrained by political interference, not only diminishes their leadership prerogatives but also hinders the implementation of strategies aimed at enhancing quality service delivery. Research in similar settings has highlighted the detrimental and pervasive impact of such practices (11,34). Additionally, poverty and lack of knowledge are prominent factors driving the community’s preference for IHFs. Literature suggests that while lower direct costs attract patients to IHFs (8), dissatisfaction with the formal healthcare system, especially for chronic conditions that require substantial psychosocial support and person-centered care, is also a significant factor (12,35–38). Improving healthcare managers’ capabilities is a key not only for enhancing access to care but also for ensuring the quality of care is centered around individual needs, thereby optimizing outcomes despite these external pressures.

### LRZ performance and functionality

During the mentorship years, a noticeable improvement was observed in both performance and functionality, compared to the baseline year of 2014. According to respondents, these enhancements were primarily driven by the improvement in leadership and strategic planning fostered through the mentorship. This aligns with findings from several other studies that highlight the positive impact of leadership enhancement on health system performance (14–16,20). Analysis of the data shows fluctuating trends, reflecting the influence of various stability-threatening events both internal and external to the HZ, as detailed by our respondents. Additionally, these fluctuations might also represent other destabilizing factors not accounted for in this study, such as sporadic security crises and disruptions in external funding, both of which heavily influence the system. These factors have been noted by other researchers as significantly affecting health outcomes in HZs (4,5,39–41). It is consistently observed that performance surpasses functionality, likely because healthcare activities tend to receive more support from external partners compared to managerial functions (42,43).

### Leadership development: The Learning Site model?

According to our respondents, the RIPSEC mentorship significantly strengthened the leadership skills of HZ managers, enhancing resource management, decision-making, and strategic planning. This progress aligns with mechanisms identified in other sub-Saharan contexts, where increased self-efficacy and perceived autonomy among managers lead to positive outcomes within supportive and conducive working environments (44,45). These findings are consistent with the realist evaluation approach, which posits that the effectiveness of support depends on specific contextual factors, actors, and mechanisms, encapsulated in the ‘Intervention-Context-Actor-Mechanism-Outcome’ (ICAMO) configuration (46,47). By fostering psychological safety and trust, RIPSEC mentorship has effectively facilitated knowledge transfer, merging theoretical insights with practical field challenges, and tailored strategies to meet the unique needs of Walungu. The importance of real-time, on-site learning for governance, as highlighted in studies from Kenya and South Africa, complements our observations of enhanced decision-making and strategic planning in Walungu (48). Similarly, the learning sites model, which supported micro-practices of governance, has provided a dynamic setting in which health managers engage in informed decision-making. This method enhances understanding and supports emergent system changes, demonstrating the value of site-based, engaged research programs, as further evidenced by a systematic review of district health system management (49).

### Sustaining achievements

Our study revealed challenges in maintaining the improvements achieved after the RIPSEC program ended. Immediately after the implementation of the RIPSEC mentorship, initial improvements in service use were observed. However, in 2018, the trend curves indicated a significant disruption in progress due to events such as the loss of skilled human resources and an epidemic emergency, as reported by our respondents. Similar constraints have been noted in other low-income contexts, where the success of interventions like RIPSEC not only relies on initial outcomes but also on the health system’s resilience to acute emergencies and the sustainability of funding and managerial practices (4,22,49,50). These findings suggest that while programs like RIPSEC can positively impact health outcomes, they are insufficient on their own to overcome all challenges. The lack of sustainable funding strategies, reliance on external aid, and instability of the health workforce could undermine the progress made in LRZ-type strengthening strategies. Experiences from other Sub-Saharan African LRZs indicate that long-term success and scalability depend on continuous engagement, adaptation of best practices, and integration of research into everyday managerial activities (22) The qualitative assessment of LRZs underscores the need to promote reflexivity in managerial decisions and to focus research on immediate LRZ challenges to foster transformative mentorship. Future studies should explore approaches that integrate these initiatives into existing healthcare systems and ensure ongoing support.

### Limits of the study

Although this study has provided valuable insights into the impact of the RIPSEC program on leadership strengthening and health service improvement in the Walungu HZ, it is not without limitations. Firstly, the retrospective design of the quantitative analysis constrains our capacity to definitively ascertain causal relationships between the RIPSEC interventions and the observed enhancements in health indicators. Secondly, the qualitative component, while providing depth through participant perspectives, may be influenced by retrospective bias or social desirability, affecting the objectivity of the responses (52). Thirdly, focusing exclusively on a single HZ limits the extent to which the findings can be generalized to other settings. Despite these considerations, the study significantly advances our understanding of the impact of health leadership development programs in contexts of crisis.

### Conclusion

This study demonstrates that the RIPSEC program positively influenced leadership enhancement and health service improvement in the Walungu HZ. Despite these gains, persistent financial challenges and the prevalent informal healthcare facilities (IHFs) pose obstacles that the program alone cannot surmount. Mentorship under RIPSEC led to better resource management, decision-making, and strategic planning, contributing to a modest increase in the utilization of formal health services. Yet, the reliance on external support and workforce instability have compromised the long-term sustainability of these improvements. This underscores the necessity for durable strategies to embed such initiatives within the existing health system framework. While the study reaffirms the critical role of leadership in public health, particularly in crisis-affected settings, it also highlights that mentorship alone is insufficient to fully address the managerial challenges HZs face. Support from external aid remains essential in crisis areas to progress towards universal health coverage.

## Data Availability

The datasets analyzed during the current study are available from the corresponding author on reasonable request.

## Aknowledgments

We extend our thanks to all those who contributed to the successful completion of this study. We are particularly grateful to the staff and management of the Walungu Health Zone for their cooperation and assistance throughout the research process. Special thanks to the RIPSEC program for their invaluable mentorship which were pivotal in conducting this research. Our gratitude also goes to staff of researchers of the ERSP-UCB for their support for this study.

